# Evaluation of Clinical Trial Data Sharing Policy in Leading Medical Journals

**DOI:** 10.1101/2020.05.07.20094656

**Authors:** Valentin Danchev, Yan Min, John Borghi, Mike Baiocchi, John P.A. Ioannidis

## Abstract

**Background:** The benefits from responsible sharing of individual-participant data (IPD) from clinical studies are well recognized, but stakeholders often disagree on how to align those benefits with privacy risks, costs, and incentives for clinical trialists and sponsors. Recently, the International Committee of Medical Journal Editors (ICMJE) required a data sharing statement (DSS) from submissions reporting clinical trials effective July 1, 2018. We set out to evaluate the implementation of the policy in three leading medical journals (JAMA, Lancet, and New England Journal of Medicine (NEJM)).

**Methods:** A MEDLINE/PubMed search of clinical trials published in the three journals between July 1, 2018 and April 4, 2020 identified 487 eligible trials (JAMA n = 112, Lancet n = 147, NEJM n = 228). Two reviewers evaluated each of the 487 articles independently. Captured outcomes were declared data availability, data type, access, conditions and reasons for data (un)availability, and funding sources.

**Findings:** 334 (68.6%, 95% confidence interval (CI), 64.1%–72.5%) articles declared data sharing, with non-industry NIH-funded trials exhibiting the highest rates of declared data sharing (88.9%, 95% CI, 80.0%–97.8) and industry-funded trials the lowest (61.3%, 95% CI, 54.3%–68.3). However, only two IPD datasets were actually deidentified and publicly available as of April 10, 2020. The remaining were supposedly accessible via request to authors (42.8%, 143/334), repository (26.6%, 89/334), and company (23.4%, 78/334). Among the 89 articles declaring to store IPD in repositories, only 17 articles (19.1%) deposited data, mostly due to embargo and regulatory approval. Embargo was set in 47.3% (158/334) of data-sharing articles, and in half of them the period exceeded 1 year or was unspecified.

**Interpretation:** Most trials published in JAMA, Lancet, and NEJM after the implementation of the ICMJE policy declared their intent to make clinical data available. However, a wide gap between declared and actual data sharing exists. To improve transparency and data reuse, journals should promote the use of unique pointers to dataset location and standardized choices for embargo periods and access requirements. All data, code, and materials used in this analysis are available on OSF at https://osf.io/s5vbg/.

## Introduction

Responsible sharing of individual-participant data (IPD) from clinical studies has gained increasing traction and has been advocated for many years by many scientists and scientific leadership organizations.^1^ However, promoting data sharing from clinical trials has not been straightforward and there has been a lot of debate surrounding privacy risks and the optimal incentives for clinical trialists and sponsors.^2-6^ Recently, the International Committee of Medical Journal Editors (ICMJE) implemented a clinical trial data sharing policy. The policy does not mandate^7,8^ data sharing but requires a data sharing statement (DSS) from submissions reporting clinical trials effective July 1, 2018.^9-11^

Prior work has identified a range of potential risks preventing trialists from sharing IPD.^3,12,13^ These risks include protection of patient privacy and confidentiality^4,13^, inappropriate data reuse and replication^12^, and researchers’ and sponsors’ potential losses of secondary publications and product advantage, respectively, due to the use of the shared data by competitors.^3,5,14^ Repositories for clinical data from industry-funded^15-17^ and publicly-funded^18^ trials have provided a safeguarded mechanism for responsible IPD sharing, thereby substantially minimizing patient-privacy and confidentiality risks. However, perceived risks of inappropriate reuse and competition have been difficult to mitigate, especially when the current reward system predominantly incentivizes high-impact publications, often based on exclusive data, at the expense of transparency, reproducibility, and data reuse.^19-22^

Disincentives for data sharing are known to have a disproportionate impact on clinical studies as the process of conducting those studies is time, cost, and labor intensive.^3^ Yet the role of prevalent disincentives and incentives (e.g., data authorship^23,24^) for clinical trial data sharing have only recently entered the public realm^3,23,25,26^, in part accelerated by discussions surrounding the ICMJE’s data sharing policy^11,27^ when many points of agreement and disagreement among stakeholders were articulated.^3,5,6,27,28^

The ICMJE policy requires investigators to state whether they will share data (or not) while simultaneously providing an opportunity for them to place multiple restrictions and conditions regarding data access. Specifically, the DSS provides an opportunity for authors and sponsors to specify periods of data exclusivity or embargo. In addition, authors can specify in the DSS how the data will be made available, reasons for data (un)availability, and related preferences. Thus, the data sharing statements, required by the ICMJE’s policy, provide a window into data sharing norms, practices, and perceived risks among trialists and sponsors.

We set out to evaluate how the ICMJE’s data sharing policy has been implemented in three leading medical journals that are also member journals of ICMJE (JAMA^9^, Lancet^10^, New England Journal of Medicine^11^ (NEJM)).

## Methods

A MEDLINE/PubMed search of clinical trials published in the three journals between July 1, 2018 and April 4, 2020 identified 629 potentially eligible articles. 486 of them included a DSS while others were either submitted before July 2018 or were letters. One article, published in 2020, met all other inclusion criteria but contained no DSS, and was included in the study sample as not sharing data on the ground that articles published in 2020 were likely submitted after July 1, 2018, and are therefore required to contain a DSS. We conducted a cross-sectional observational study for all 487 article (JAMA n = 112, Lancet n = 147, NEJM n = 228). Two reviewers evaluated each article independently. Discrepancies were resolved unanimously or by a third reviewer.

Data was classified as available when authors answered “Yes” (JAMA, NEJM) or gave an unstructured positive response (Lancet) to the data availability question. Information about data type, access, conditions and reasons for data (un)availability were taken from the DSS. We also compared declared to actual data availability in repositories by examining whether information about data and data themselves are available in the respective repository.

Funding sources were classified as industry, non-industry NIH, non-industry non-NIH, and mixed. Industry refers to research funding from companies. Non-industry NIH refers to research funding from the U.S. National Institutes of Health (NIH). Non-industry non-NIH refers to research funding from foundations, trusts, associations, national institutes outside the USA, etc. Mixed refers to any combination of the other research-funding categories.

We conduct descriptive analysis of variables associated with data sharing by type of funding and publication journal. For the primary outcome variable, declared data sharing, we report the 95% confidence intervals determined by bootstrapping (100,000 iterations). We used Python programming language (Python Software Foundation, available at https://www.python.org/) and Jupyter Notebook to perform data analysis and to generate summary statistics and graphs. All computer code used for this analysis is referenced in Appendix 1 and is available for download and reuse at https://osf.io/s5vbg/.

For details on the MEDLINE/PubMed search strategy, inclusion and exclusion criteria for the study, definition of variables, and data extraction, see Supplementary Material.

## Results

Overall, 334 (68.6%, 95% confidence interval (CI), 64.1%–72.5%) articles declared data sharing (Table 1). Prevalence of declared data sharing varied by journal and funder type. Non-industry NIH-funded trials had the highest rates of declared data sharing (88.9%, 95% CI, 80.0%–97.8) and industry-funded trials the lowest (61.3%, 95% CI, 54.3%–68.3) (Figure 1A). The highest rate of declared data sharing of NIH-funded trials is consistent across the three journals (Figure 1B). No substantial changes in the prevalence of declared data sharing were observed over the span of the first seven quarters of policy implementation (Figure 1C).

**Figure 1.**
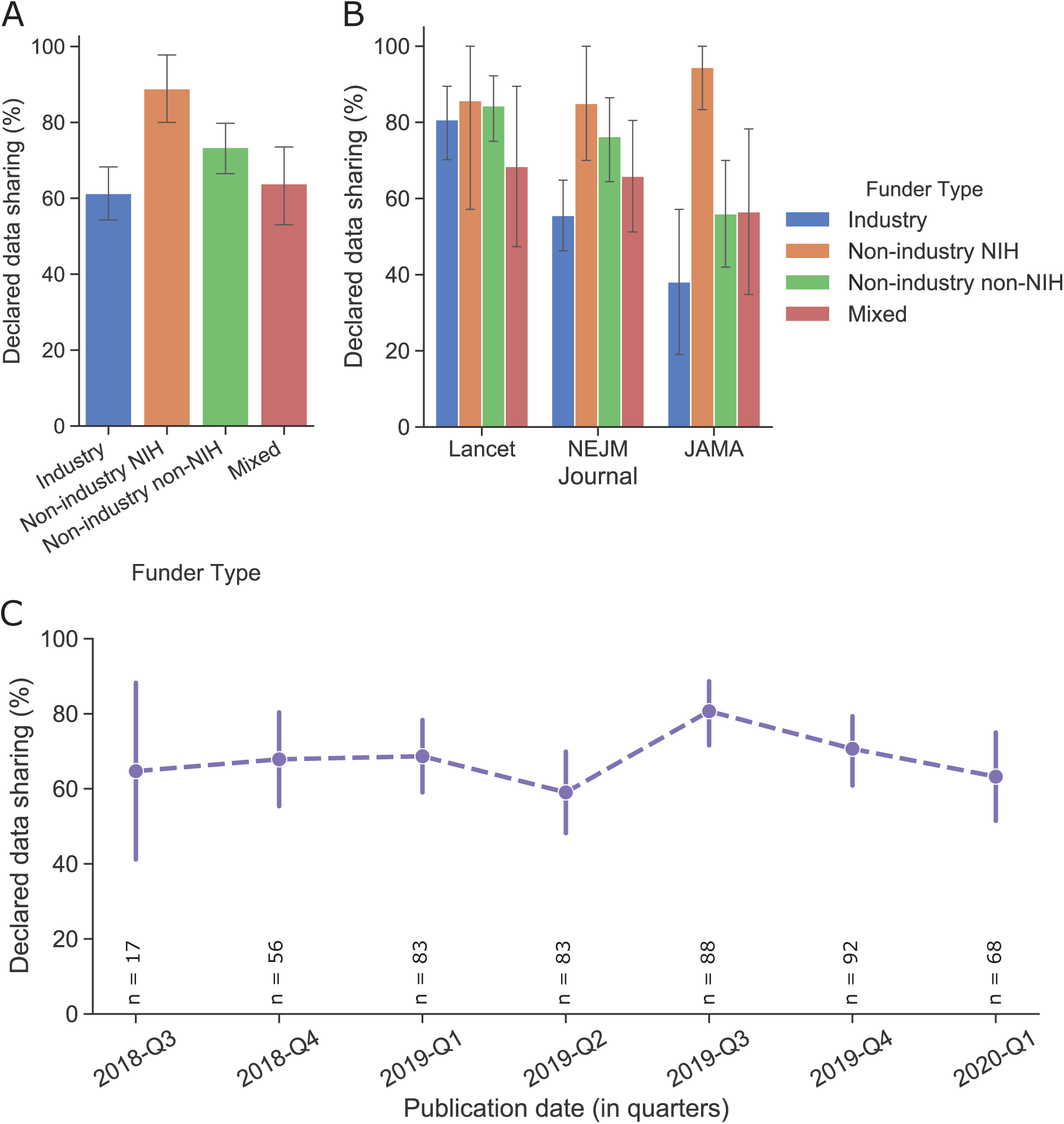
Declared clinical trial data sharing in leading medical journals. Percentage of articles that declared their intend to share clinical trial data by (A) Funder type, (B) Funder type and Journal, (C) Publication date represented in quarters. Error bars represent 95% confidence intervals computed via bootstrapping, 100,000 iterations. *N* = 487 articles.

**Table 1.** Prevalence and Conditions of Declared Clinical Trial Data Sharing by Type of Funding

|  |  |  |  |  |  |
| --- | --- | --- | --- | --- | --- |
|  | **Funder Type** |  |  |  |  |
|  | **Industry (n = 186)** | **Non-industry NIH (n = 45)** | **Non-industry non-NIH (n = 173)** | **Mixed (n = 83)** | **Total (n = 487)** |
|  | **No./Total (%)** | **No./Total (%)** | **No./Total (%)** | **No./Total (%)** | **No./Total (%)** |
| **Declared data sharing<sup>a</sup>** |  |  |  |  |  |
| Yes | 114 (61.3) | 40 (88.9) | 127 (73.4) | 53 (63.9) | 334 (68.6) |
| No | 72 (38.7) | 5 (11.1) | 46 (26.6) | 30 (36.1) | 153 (31.4) |
| **Journal** |  |  |  |  |  |
| JAMA | 21 (11.3) | 18 (40.0) | 50 (28.9) | 23 (27.7) | 112 (23.0) |
| Lancet | 57 (30.6) | 7 (15.6) | 64 (37.0) | 19 (22.9) | 147 (30.2) |
| NEJM | 108 (58.1) | 20 (44.4) | 59 (34.1) | 41 (49.4) | 228 (46.8) |
| **Type of declared available data<sup>b</sup>** |  |  |  |  |  |
| Deidentified individual participant data | 83/114 (72.8) | 33/40 (82.5) | 95/127 (74.8) | 44/53 (83.0) | 255/334 (76.3) |
| Aggregate data only | 2/114 (1.8) | 0/40 (0.0) | 1/127 (0.8) | 0/53 (0.0) | 3/334 (0.9) |
| Unspecified/ partial data<sup>c</sup> | 29/114 (25.4) | 7/40 (17.5) | 31/127 (24.4) | 9/53 (17.0) | 76/334 (22.8) |
| **Access to data<sup>d</sup>** |  |  |  |  |  |
| Request to authors | 9/114 (7.9) | 15/40 (37.5) | 96/127 (75.6) | 23/53 (43.4) | 143/334 (42.8) |
| Request to committee/ group/ unit | 15/114 (13.2) | 3/40 (7.5) | 36/127 (28.3) | 13/53 (24.5) | 67/334 (20.1) |
| Request to repository/ archive | 46/114 (40.4) | 22/40 (55.0) | 5/127 (3.9) | 16/53 (30.2) | 89/334 (26.6) |
| Request to company | 72/114 (63.2) | 0/40 (0.0) | 0/127 (0.0) | 6/53 (11.3) | 78/334 (23.4) |
| Access unspecified | 5/114 (4.4) | 2/40 (5.0) | 5/127 (3.9) | 3/53 (5.7) | 15/334 (4.5) |
| Data is available to others | 0/114 (0.0) | 0/40 (0.0) | 2/127 (1.6) | 0/53 (0.0) | 2/334 (0.6) |
| **Conditional data access<sup>d</sup>** |  |  |  |  |  |
| Data embargo | 49/114 (43.0) | 22/40 (55.0) | 57/127 (44.9) | 30/53 (56.6) | 158/334 (47.3) |
| Up to 1 year<sup>c</sup> | 20/114 (17.5) | 13/40 (32.5) | 30/127 (23.6) | 18/53 (34.0) | 81/334 (24.3) |
| Above 1 year to 2 years | 13/114 (11.4) | 7/40 (17.5) | 13/127 (10.2) | 6/53 (11.3) | 39/334 (11.7) |
| Above 2 years | 4/114 (3.5) | 2/40 (5.0) | 4/127 (3.1) | 5/53 (9.4) | 15/334 (4.5) |
| Product approval | 36/114 (31.6) | 0/40 (0.0) | 0/127 (0.0) | 1/53 (1.9) | 37/334 (11.1) |
| Collaboration | 0/114 (0.0) | 2/40 (5.0) | 6/127 (4.7) | 1/53 (1.9) | 9/334 (2.7) |
| **Reasons for why data not available<sup>d</sup>** |  |  |  |  |  |
| No reason given | 40/72 (55.6) | 2/5 (40.0) | 26/46 (56.5) | 15/30 (50.0) | 83/153 (54.2) |
| Data privacy | 3/72 (4.2) | 1/5 (20.0) | 9/46 (19.6) | 2/30 (6.7) | 15/153 (9.8) |
| Time and cost | 0/72 (0.0) | 0/5 (0.0) | 2/46 (4.3) | 0/30 (0.0) | 2/153 (1.3) |
| Ongoing trial/ research | 2/72 (2.8) | 0/5 (0.0) | 2/46 (4.3) | 9/30 (30.0) | 13/153 (8.5) |
| Regulatory approval | 3/72 (4.2) | 0/5 (0.0) | 1/46 (2.2) | 0/30 (0.0) | 4/153 (2.6) |
| Proprietary data | 7/72 (9.7) | 0/5 (0.0) | 4/46 (8.7) | 3/30 (10.0) | 14/153 (9.2) |
| Shared among co-investigators only | 2/72 (2.8) | 0/5 (0.0) | 0/46 (0.0) | 0/30 (0.0) | 2/153 (1.3) |
| Data may be available for collaboration | 6/72 (8.3) | 0/5 (0.0) | 0/46 (0.0) | 1/30 (3.3) | 7/153 (4.6) |
| Data may be available upon request | 19/72 (26.4) | 2/5 (40.0) | 6/46 (13.0) | 7/30 (23.3) | 34/153 (22.2) |

<sup>a</sup>The denominators represent the total for the respected column, unless otherwise indicated. <sup>b</sup>Detailed definitions of variables are provided in the Supplementary Material and in the Data dictionary. <sup>c</sup>Among the articles not specifying the type of data, 13 intended to store data in clinical trial data repositories, presumably individual participant data, and one made available (via collaboration) individual participant data in a designated repository.
<sup>d</sup>Categories are not mutually exclusive. <sup>e</sup>Time periods are not specified in all articles proposing data embargo.

Regarding type of shared data, 76.3% (255/334) of the articles proposed to provide deidentified IPD, 22.8% (76/334) unspecified, and 0.9% (3/334) aggregate data only. Only two IPD datasets were actually deidentified and publicly available (on journal website) as of April 10, 2020 (both datasets were associated with the same clinical trial). The remaining were supposedly accessible via request to authors (42.8%), repository (26.6%), and company (23.4% overall, 63.2% among industry-funded trials).

Conditions for access to data included embargo (47.3%, 158/334), product approval (11.1%, 37/334), and collaboration (2.7%, 9/334). Among the 158 articles specifying embargo, about half required one year or less of data exclusivity. In the other half of embargo cases, the embargo period exceeded 1 year or was unspecified.

Data repositories have a central role in improving sharing, security, discoverability, and reuse of research data,^29,30^ and in particular of individual-level participant data from clinical trials.^31-33^ Among the 89 articles proposing to make IPD available through repositories, many planned to store data in general-purpose repositories, including the Clinical Study Data Request (n = 31), the Yale Open Data Access (YODA) Project (n = 7), and Vivli (n = 7). Another 30 articles planned to store IPD in NIH-supported domain-specific data repositories such as NCTN/NCORP Data Archive (n = 10), the NHLBI Biologic Specimen and Data Repository Information Coordinating Center (BioLINCC) (n = 9), and the NICHD Data and Specimen Hub (DASH) (n = 5) (Figure 2 and Table S1).

**Figure 2.**
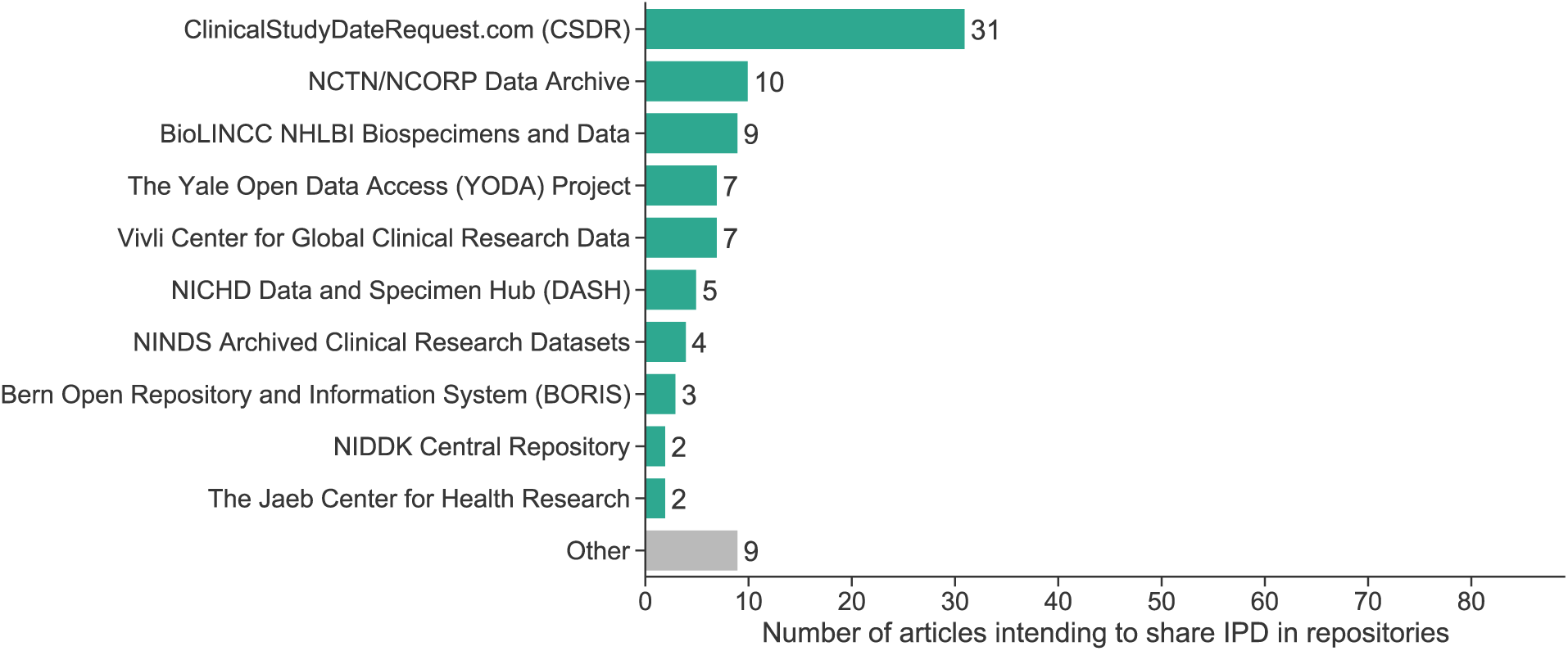
Ranking of data repositories by the number of articles intending to share individual participant data (IPD) in the respective repository. Repositories referenced in only one article/DSS are aggregated into category Other. Acronyms: *NCTN* National Clinical Trials Network; *NCORP* National Cancer Institute (NCI) Community Oncology Research Program; *BioLINCC* Biologic Specimen and Data Repository Information Coordinating Center; *NHLBI* National Heart, Lung, and Blood Institute; *NICHD* Eunice Kennedy Shriver National Institute of Child Health and Human; *NINDS* National Institute of Neurological Disorders and Stroke; *NIDDK* National Institute of Diabetes and Digestive and Kidney Diseases. See Table S1 for details about the clinical trial data repositories.

We compared declared to actual data availability in repositories (Table 2). Among 89 articles, information about the data was uncommon to find in the repository (22.5%, 20/89) and the data themselves were even less frequently available there (19.1%, 17/89). Although data of NIH-funded trials (31.8%) were somewhat more likely than data of industry-funded trials (15.2%) to be available in repositories, most trials provided neither information nor data in the respective repositories, mostly due to embargo and pending regulatory approval. Specifically, among the 72 articles that declared their intent but did not store data on repository, 37 (51.4%) made data access conditional on embargo or product approval.

**Table 2.**
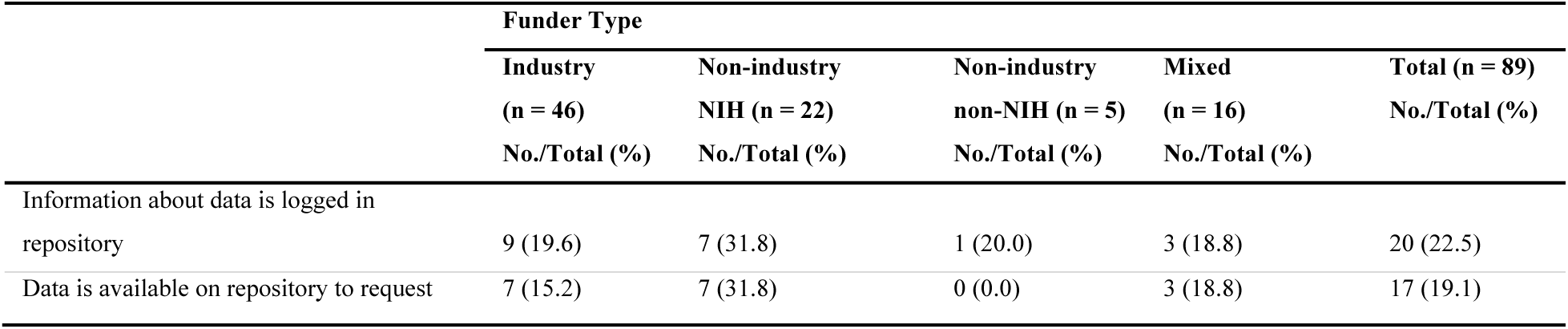
Declared versus Actual Availability of IPD in Repository by Type of Funding

Among reasons for data withholding, articles referred to data privacy (9.8%, 15/153), proprietary data (9.2%, 14/153), ongoing research (8.5%, 13/153), and pending regulatory approval (2.6%, 4/153). Most articles withholding data (54.2%, 83/153) provided no reason. One article, which had no intent to make data available to others, was retracted.

## Discussion

Most trials published in JAMA, Lancet, and NEJM after the endorsement of the ICMJE policy declared their intent to make clinical data available. Non-industry funded trials communicated greater intent to share data than industry-funded trials. However, the commitment to data sharing substantially decreases when we consider indicators of actual versus declared data sharing—out of 334 articles declaring to share data, only two IPD datasets were actually deidentified and publicly available on journal website, and among the 89 articles declaring to store IPD in repositories, data from only 17 articles were found on the respective repository (Figure 3).

**Figure 3.**
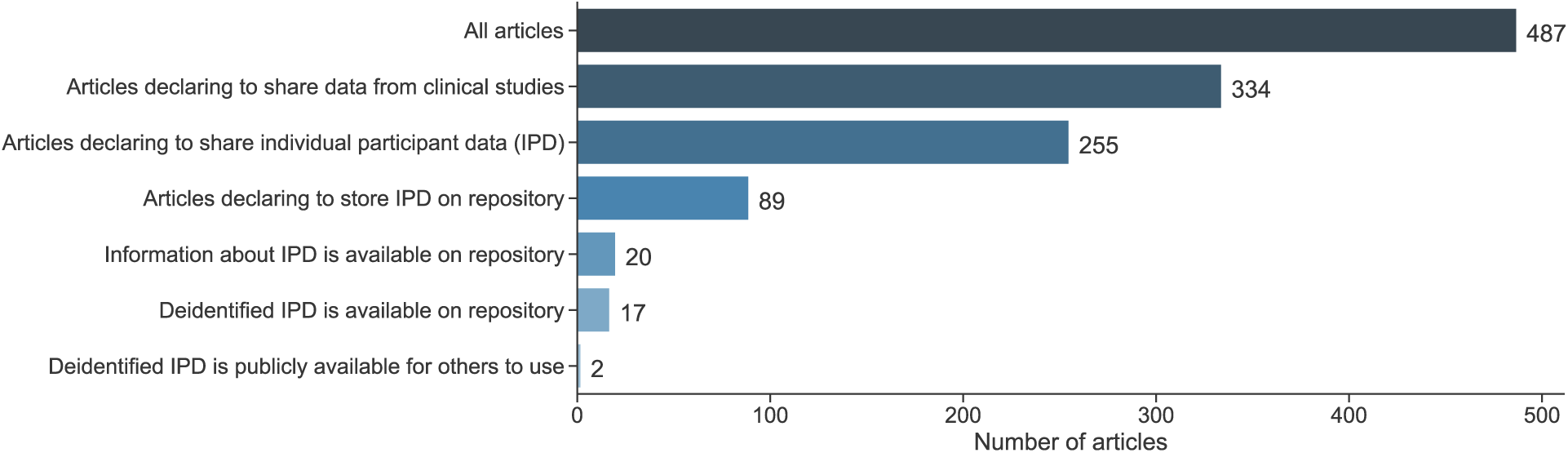
Indicators of declared versus actual clinical trial data sharing.

Consistent with prior research of clinical trial data registries^34,35^ and data sharing statements,^7^ DSS language was often ambivalent. Offering of aggregate data, collaboration demands, lengthy or unspecified embargo periods, and the use of legacy methods for access such as author or company request communicate only lukewarm commitment. Repositories can be instrumental for sharing but real practices may diverge from intent.

Major funders are in the midst of designing or updating data sharing policies. Recently, for example, the National Institutes of Health (NIH) has drafted and requested public comments on a data sharing policy.^25^ The draft data sharing policy was discussed in the context of clinical trial data sharing.^21^ Our findings highlighted inefficiencies in clinical trial data sharing practices, and addressing those in NIH and other major funders’ policies could narrow the wide gap we identified between declared and actual availability of clinical trial IPD.

Our study has limitations that should be acknowledged. First, only three journals were considered. Moreover, we could readily investigate declared versus actual data sharing practices only for repositories. Finally, only two IPD datasets were deidentified and available on journal website, so we could not meaningfully examine the usability of shared data or reproducibility^8^ of the clinical trial studies. As more IPD datasets become available, it would be interesting to assess whether they are easy to use, and how complete is the information being provided.

To promote transparency and data reuse, journals and funders should work towards incentivizing data sharing via funding mechanisms^21^ and data authorship^23^, and simultaneously discourage ambivalent wording in DSS and possibly mandate data sharing. They can promote the use of unique pointers to dataset location in repositories and to data request forms. Standardized choices for embargo periods, access requirements, and conditions for data use as part of the data sharing process could also reduce unnecessary data withholding and turn declarative data sharing into actual transparency in clinical trial data.

## Data Availability

All the data, computer code, and materials for this study are publicly available from the Open Science Framework at https://osf.io/s5vbg/.

## Funding/Support

METRICS is supported by a grant from the Laura and John Arnold Foundation.

## Role of the Funder/Sponsor

The funder had no role in the design, data collection, analysis, and interpretation of data, or preparation, review and approval of the manuscript, or decision to submit the manuscript for publication.

## Supplementary Material

### Appendix 1: Inclusion criteria, search strategy, and data collection and analysis

#### Inclusion and exclusion criteria

In order to be eligible to be selected in the study, a published paper must meet the following inclusion criteria:

1. Publication reports clinical trial results
2. Published in JAMA, NEJM, Lancet
3. Published since July 1, 2018
4. Type of publication is Article
5. Contain a Data Sharing Statement (DSS)*

*Articles published in 2020 that meet criteria 1 to 4 are eligible even if no DSS is present as these are likely submitted after July 1, 2018, and therefore fall under the requirements of the ICMJE data sharing policy.

Excluded from the study were publications that contain no Data Sharing Statement due to:

1. Submission prior to July 1, 2018
2. Study not a clinical trial (e.g., observational)
3. Type of publication is letter/correspondence

#### Search strategy

A MEDLINE/PubMed search was performed on April 4, 2020 using the following search strategy:

~~~
(((("The New England journal of medicine"[Journal]) OR "JAMA"[Journal]) OR "Lancet (London, England)"[Journal]) AND clinical trial[Publication Type]) AND ("2018/07/01"[Date - Publication] : "2020/04/04"[Date - Publication])
~~~

As of April 04 2020, the search yielded 629 results. Out of those, 486 publications were clinical trials and contained a DSS. One 2020 publication met 1 to 4 inclusion criteria but contained no DSS, and was included in the study sample as not sharing data because articles published in 2020 were likely submitted after policy’s effective date, July 1, 2018. We conducted a cross-sectional observational study of the resulting sample of 487 articles.

#### Independent review of articles

For each of the 487 articles, two reviewers independently evaluated the Data Sharing Statement and funding statement, using procedures described in the Codebook. Discrepancies were resolved unanimously or by a third reviewer.

#### Data analysis

We performed data analysis and generated summary statistics and data visualizations using Python (Python Software Foundation. Python Language Reference, version 3.6.8. Available at http://www.python.org), `Jupyter Notebook`,^1^ and the following libraries: `SciPy`,^2^ `Pandas`,^3^ `NumPy`,^4^ `Matplotlib`,^5^ `Scikits-Bootstrap`,^6^ `Seaborn`.^7^ All computer code used in this analysis is available at https://osf.io/s5vbg/.

### Appendix 2: Codebook

#### A. Declared data sharing

1 = YES → Code **B**, **C**, **D**, and **F**

0 = NO → Code **E** and **F**

#### B. Type of declared available data [1 if applicable, blank otherwise]

Deidentified individual participant data

Aggregate data only

Unspecified/ partial data

#### C. Access to data [1 if applicable, blank otherwise]

Request to authors

Request to committee/ group/ unit

Request to company

Request to repository/ archive

Name of repository [Name as noted on DSS]
Repository contains information about the data/study [1 = Yes, 0 = No]
Data is available on repository [1 = Yes, 0 = No]

Access unspecified

Data is available to others

#### D. Conditions for data sharing [1 if applicable, blank otherwise]

Embargo

Embargo period [in Months]
If embargo period unspecified, copy wording from the DSS

Collaboration

Product approval

#### E. Reasons for why data not available [1 if applicable, blank otherwise]

No reason given

Data privacy

Time and cost

Ongoing trial/ research

Regulatory approval

Proprietary data

Shared among co-investigators only

Data may be available for collaboration

Data may be available upon request

#### F. Funder type*

1 = Industry

2 = Non-industry NIH

3 = Non-industry non-NIH

4 = Mixed (any combination of industry, non-industry NIH, non-industry non-NIH)

* Only funding is eligible (in-kind support or supplies provided at no charge are not eligible).

### Appendix 3: Detailed description of clinical trial data repositories

**Table S1.** Description of Clinical Trial IPD Repositories^a^

| Table S1. Description of Clinical Trial IPD Repositories<sup>a</sup> |  |  |  |
| --- | --- | --- | --- |
| Acronym / short name | Full name | URL | Host organization |
| CSDR | Clinical Study Data Request | https://www.clinicalstudydatarequest.com | Consortium of clinical study sponsors/funders |
| NCTN/NCORP Data Archive | National Clinical Trials Network (NCTN) and NCI Community Oncology Research Program (NCORP) Data Archive | https://nctn-data-archive.nci.nih.gov | National Cancer Institute (NCI) |
| BioLINCC | Biologic Specimen and Data Repository Information Coordinating Center | https://biolincc.nhlbi.nih.gov | National Heart, Lung, and Blood Institute (NHLBI) |
| YODA | The Yale University Open Data Access (YODA) Project | https://yoda.yale.edu/ | Yale University |
| Vivli | Center for Global Clinical Research Data | https://vivli.org | The Multi-Regional Clinical Trials Center of Brigham and Women's Hospital and Harvard |
| DASH | NICHHD Data and Specimen Hub | https://dash.nichd.nih.gov/ | The Eunice Kennedy Shriver National Institute of Child Health and Human Development (NICHD) |
| NINDS Archived Clinical Research Datasets | NINDS Archived Clinical Research Datasets | https://www.ninds.nih.gov/Current-Research/Research-Funded-NINDS/Clinical-Research/Archived-Clinical-Research-Datasets | National Institute of Neurological Disorders and Stroke (NINDS) |
| BORIS | Bern Open Repository and Information System | https://boris.unibe.ch/ | University of Bern and the Bern University Hospital |
| NIDDK Central Repository | NIDDK Central Repository | https://repository.niddk.nih.gov | National Institute of Diabetes and Digestive and Kidney Diseases (NIDDK) |
| JCHR | JAEB Center for Health Research | https://www.jaeb.org/ | JAEB Center for Health Research |

<sup>a</sup> Included are only repositories that were referenced in two or more data sharing statements.

## Notes

### Competing Interest Statement

The authors have declared no competing interest.

